# The Refractive Error and Vision Impairment Estimation With Spectacle data (REVIEWS) study

**DOI:** 10.1101/2021.06.15.21258945

**Authors:** Michael Moore, James Loughman, John S. Butler, Arne Ohlendorf, Siegfried Wahl, Daniel I. Flitcroft

## Abstract

**Objective:** To investigate whether spectacle lens sales data can be used to estimate the population distribution of refractive error amongst ametropes and hence estimate the current and future risk of vision impairment.

**Design:** Cross Sectional Study

**Subjects:** A total 141,547,436 spectacle lens sales records from an international European lens manufacturer between the years 1998 and 2016.

**Methods:** Anonymized patient spectacle lens sales data including refractive error information was provided by a major European spectacle lens manufacturer. Data from the Gutenberg Health Survey was digitized to allow comparison of a representative, population-based sample to the spectacle lens sales data. A bootstrap analysis was completed to assess the comparability of both datasets. The expected level of vision impairment due to myopia at age 75 was calculated for both datasets using a previously published risk estimation equation combined with a saturation function.

**Main Outcome Measures:** Comparability of spectacle lens sales data on refractive error to typical population surveys of refractive error and its potential utility to predict vision impairment due to refractive error.

**Results:** Equivalent estimates of the population distribution of spherical equivalent refraction can be provided from spectacle lens data within limits. For myopia, the population distribution was equivalent to the Gutenberg Health Survey (≤ 5% deviation) for levels ≤-2.0 dioptres, while for hyperopia the distribution was equivalent (≤ 5% deviation) for levels ≥ +3.0 diopters. The estimated rates of vision impairment due to myopia were not statistically significantly different (χ^2^ = 182, DoF = 169, p = 0.234) between the spectacle lens data and Gutenberg Health Survey data.

**Conclusions:** The distribution of refractive error and hence the risk of vision impairment due to refractive error within a population can be determined using spectacle lens sales data. Pooling this type of data from multiple industry sources could provide a cost effective, timely and globally representative mechanism for monitoring the evolving epidemiology of refractive error and associated vision impairment.

## Introduction

Vision impairment is a huge challenge internationally which is projected to worsen as a consequence of global population aging unless significant effort is made to address the many underlying causes. Refractive error has been identified as a risk factor for the development of numerous ocular pathologies which can lead to vision impairment. Significant refractive errors, both myopic and hyperopic, are known to be amblyogenic in children.^1^ Higher degrees of hyperopia is a risk factor for the development of age-related macular degeneration (AMD),^2^ while higher levels of myopia are known to increase the risk of glaucoma,^3^ cataract,^4^ retinal detachment^5^ and myopic maculopathy.^6^

The individual and societal cost of vision impairment is substantial. Societal costs can be measured by the loss of productivity^7^ and the need to provide adequate medical care and support to those affected by vision impairment.^8^ Those with vision impairment are more likely to require support in day to day living, suffer from falls and have health or emotional problems interfere with their life.^8,9^ Quality of life is also significantly affected, with vision impairment having a similar impact as stroke, heart attack and diabetes,^10^ and even mild vision impairment associated with reduced quality of life.^11^

Refractive error typically develops in childhood.^12^ The association between refractive error and vision impairment, however, does not become apparent for many decades and is a function of refractive error type and magnitude as well as increasing age.^13–15^ Myopia is the refractive error that is of most concern. It has been demonstrated that there is an increased lifetime risk of vision impairment with all levels of myopia, but particularly at higher levels.^13^ A recent meta-analysis indicated that one in three high myopes are at risk of bilateral vision impairment within their lifetime and that even low to moderate myopes are at significantly increased risk of ocular disease and disability.^16^ There is an increasing amount of evidence that the prevalence of myopia within the population has increased over the last number of decades. The most significant increases have been observed in Asian populations^17^ with some countries seeing over 90% of children become myopic by the late teenage years.^18,19^ Although ethnicity appears to play a role, there is evidence of increasing prevalence of myopia in many populations around the world.^20–23^

It is important to have current and easily accessible refractive error epidemiological data in order to plan appropriate public health resource allocation to meet the need for correction of refractive error and treatment of any associated pathology, particularly in the context of a changing population burden of refractive error. Holden et al.’s landmark paper^24^ predicted that by 2050 almost 50% of the global population will be myopic, with nearly 10% of the population falling into the highly myopic category (using a threshold of -5 D). This is of great concern given the likelihood of increased levels of vision impairment due to both uncorrected refractive error and the ocular pathology associated with myopia. Holden et al.^24^ used existing epidemiological studies to make their predictions. They identified the lack of epidemiological data in “many countries and age groups, across representative geographic areas”^24^ as a significant limitation of their study, with predictions of high myopia prevalence particularly susceptible to the paucity of available evidence. The lack of epidemiolocal data is not surprising given the time and financial investment required to carry out these studies.

As the risk of vision loss associated with increasing refractive error is non-linear,^13,15,16^ it is not sufficient to merely establish the proportion of the population affected by myopia or hyperopia. It is necessary to determine the number of individuals affected by different levels of refractive error within a population to gain a true insight into the population risk of vision impairment due to refractive error.

Spectacle lens sales data represents a potential source of contemporary refractive error data which, if made accessible, could provide valuable insights into the changing epidemiology of refractive error and associated risks of vision impairment. The value and limitations of spectacle lens sales data as an epidemiological tool to determine refractive error distribution in a population has previously been described.^25^ Principally, the distribution of refractive error found in spectacle lens sales data does not follow standard population distributions of refractive error as individuals with no refractive error do not typically purchase spectacles lenses, hence emmetropes and near-emmetropes are under-represented in such data. The symptomatic nature of higher levels of refractive error implies that the majority of the population affected are likely to use spectacles, particularly in high income countries where the visual demands associated with education and employment are high and where subsidised access to eyecare is available.^26^ Most studies of refractive error epidemiology report their distributions across the entire range of refractive error. By concentrating analysis on the myopic and hyperopic ends or tails of the distribution, rather than the central emmetropic range of the distribution it may be possible to use spectacle lens sales data as an epidemiological tool.

The aim of this paper, therefore, was to investigate whether spectacle lens sales data can be used to estimate the population distribution of refractive error amongst ametropes and hence estimate the current and future impact of refractive errors on the risk of vision impairment.

## Methods

Anonymized patient spectacle lens sales data were provided by a major European spectacle lens manufacturer. This dataset (n=141,547,436) comprised lenses that had been manufactured and dispatched after an order was received from an eye care practitioner, with the majority (> 98%) of lenses for delivery within Europe. The data was collated into histogram data using the SQLite database engine (Hipp, Wyrick & Company, Inc., Charlotte, North Carolina, USA) and analyzed using the R statistical programming language (R Core Team (2021). R: A language and environment for statistical computing. R Foundation for Statistical Computing, Vienna, Austria. URL https://www.R-project.org/.). The data provided included the spherical power, cylindrical power and axis of the spectacle prescription, lens design, diameter, laterality (prescribed for right or left eye) and date of manufacture. For lens designs with an addition, this was also specified. The presence of an addition allowed the lenses to be separated into two groups, the single vision (SV) lens group and the addition (ADD) lens group. The data was validated for missing and malformed data fields and any lenses with incomplete or invalid data were excluded. The spherical equivalent power was calculated for each lens.

Data from the Gutenberg Health Study^27^ (GHS) study was extracted by digitizing the published results using Plot Digitiser (http://plotdigitizer.sourceforge.net/). The GHS was chosen as a comparison for several reasons. Firstly, the GHS took place in Mainz, Germany and the manufacturer database reflected almost exclusively European lens sales, with Germany the largest contributor (≈ 48%). Secondly, as the spectacle lens data comprised a substantial proportion of reading addition lenses typically used by older presbyopic adults^28^ (age ≥ 40-45 typically),^29^ the adult age profile of the GHS (age 35-74 years) was comparable.

Myopia and hyperopia were analyzed using the definitions given by the GHS i.e., a spherical equivalent (SE) refractive error of < -0.50 D being considered myopic and a SE refractive error of > 0.75 D being considered hyperopic. High myopia was defined as SE ≤ -6.00 D. The International Myopia Institute recommends the adoption of an agreed standard for myopia of ≤ -0.50 D^30^ so results using this criterion are also reported.

To determine confidence intervals of the estimates, a bootstrapping technique was used to generate 1,000 new distributions of refractive error from the SV and ADD lens data, with each new distribution comprising the same original sample size as the GHS (n = 13,959). The new distributions were generated by random sampling with replacement using the ‘infer’ extension package for R. With this technique, the mean number of cases for each 1 D bin value of spherical equivalent was calculated along with 95% confidence intervals. This was repeated for both the myopic and hyperopic tails of the distributions and the results were compared to the GHS distribution of refractive error.

In order to determine the range of refractive error values over which the bootstrap analysis should take place, the analysis was repeated with different spherical equivalent starting values, starting at 0 D spherical equivalent and changing in 1 D steps for both the myopic and hyperopic tails of the distribution. This allowed the deviance between the calculated 95% confidence intervals and the GHS distribution to be determined.

The final fitted bootstrapped distributions were generated which allowed comparison with the GHS. The proportion of each diopter value of myopia and hyperopia was calculated within the range that was found to match the GHS well. The expected level of vision impairment due to myopia at each diopter value of myopia was determined using equation 1 as described by Bullimore et al^31^ and was modelled on published data and models that relate refractive error and age to vision impairment risk.^13,15^ A saturation function (equation 2) was applied to this equation to avoid overestimation of vision impairment at higher levels of myopia. This allowed the expected level of vision impairment by age 75 for a sample of 100,000 SV spectacle lens users to be calculated. This was compared to the expected level of vision impairment at age 75 over the same range of myopia for participants in the GHS. 

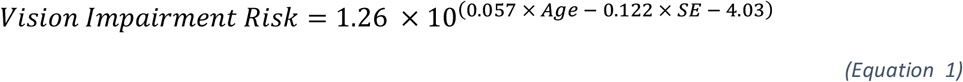

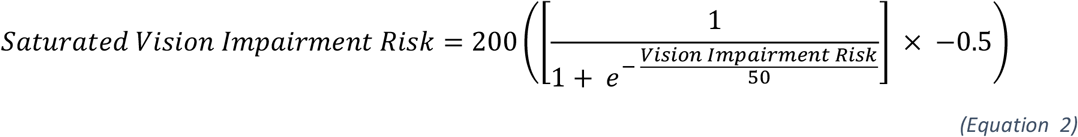

## Results

The spectacle lens dataset comprised 141,547,436 lenses from the manufacturer sales records ranging from the year 1998 to 2016. Records with incomplete or missing data were excluded, and only years with complete data were included in the analysis. In total 134,280,063 spectacle lenses were included, comprised of 84,561,994 SV lenses and 49,709,191 ADD lenses.

The distribution of refractive error for the SV, ADD lenses and the GHS are shown in Figure 1. All distributions demonstrate the classic negatively skewed leptokurtotic curve found in most studies of refractive error, with the majority of observations centred close to emmetropia. The only exception to this pattern was the SV spectacle lenses which were found to have a bimodal distribution with a significant notch apparent at zero spherical equivalent. Table 1 shows the proportion of myopia and hyperopia found in each dataset. The most significant difference observed was in the proportion of emmetropia present, with much lower levels in all spectacle lens datasets.

**Table 1:** Proportion of refractive error types in each dataset.

| | Total Emmetropia (SE $\geq -0.50$ D and $\leq +0.75$ D) | Total Hyperopia (SE $> +0.75$ D) | Total Myopia (SE $< -0.50$ D) | Total Myopia (SE $\leq -0.50$ D) | High Myopia (SE $\leq -6.00$ D) |
| --- | --- | --- | --- | --- | --- |
| All Spectacle Lenses | 19.1% | 44.9% | 36.0% | 38.0% | 4.8% |
| SV Lenses | 15.5% | 44.9% | 39.6% | 41.7% | 5.3% |
| ADD Lenses | 25.0% | 45.1% | 29.9% | 31.9% | 4.0% |
| GHS | 35.1% | 29.8% | 35.1% | 39.9% | 3.5% |

**Figure 1:**
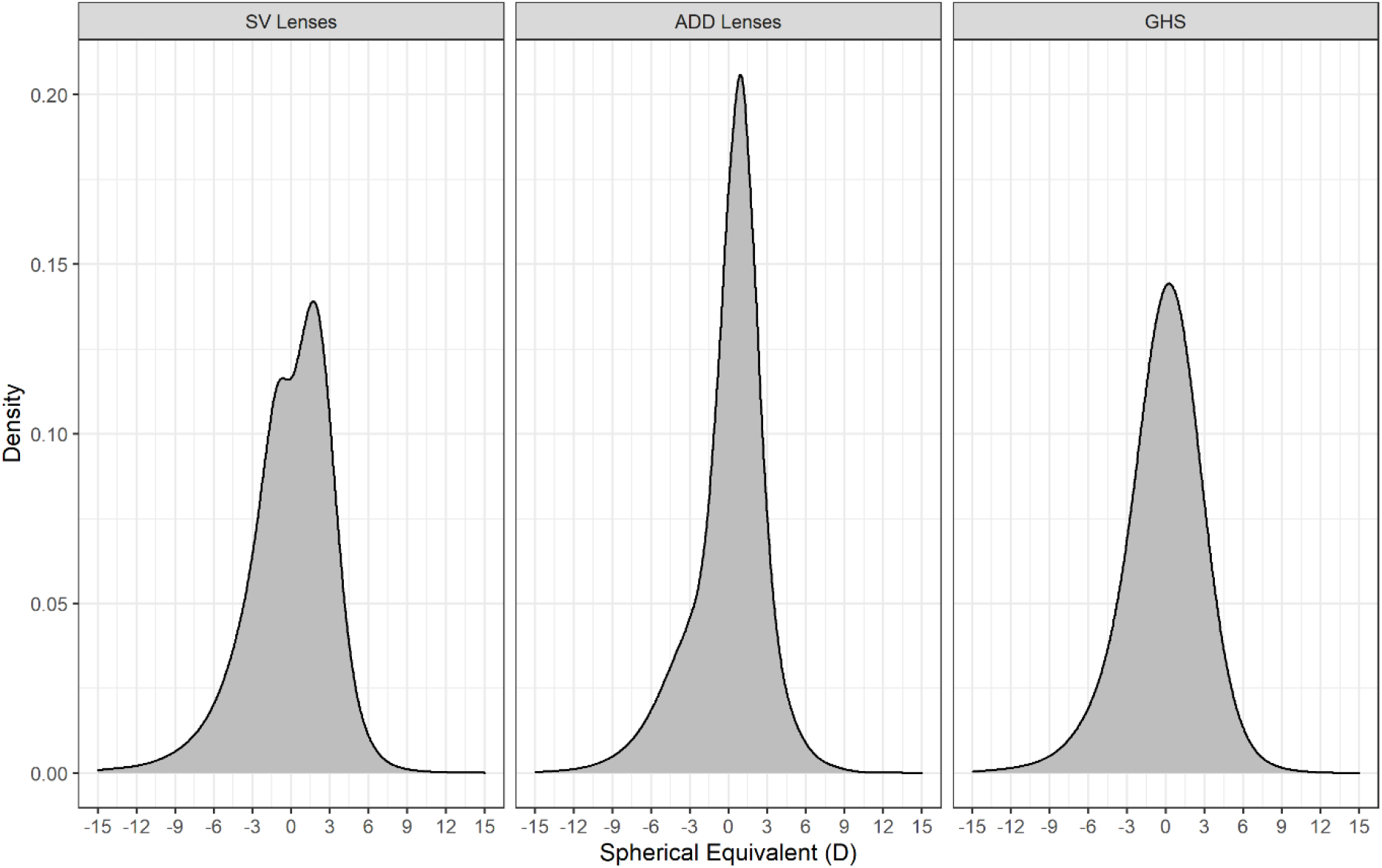
Spectacle Lens Distribution of refractive error from manufacturer data for single vision (SV) lenses (n = 84,561,994), addition (ADD) lenses (n = 49,709,191) and from the Gutenberg Health Survey (n= 13,959).

Repeating the bootstrapping technique for both the myopic and hyperopic tails of each distribution, it was found that the deviation between the actual occurrence of each 1 D value of spherical equivalent for the GHS and all of the spectacle lens data sets was greatest (over 50%) at 0 D spherical equivalent. The deviation reduced to 5% or less between -2 D and -15 D for the myopic end of the distributions and between +3 D and +15 D for the hyperopic end of the distributions (Figure 2).

**Figure 2:**
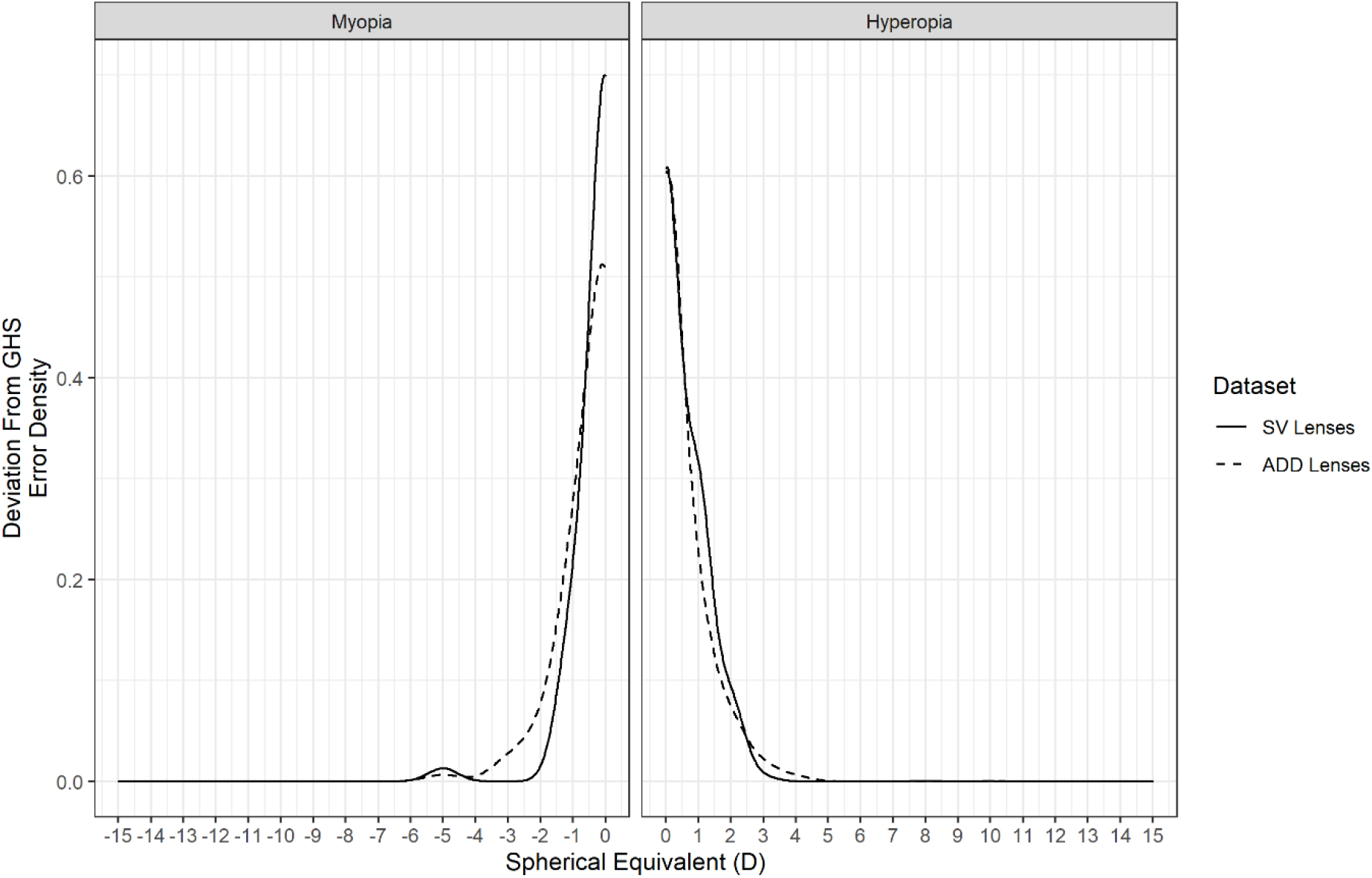
Deviation between bootstrapped confidence intervals and the observed occurrence of refractive error in the Gutenberg Health Survey. The deviation is greatest when starting at zero diopters spherical equivalent and trends towards zero at higher absolute values of spherical equivalent.

Figures 3 and 4 show the mean number of lenses with 95% confidence intervals for each 1 D from all 1,000 generated distributions for the myopic and hyperopic tails of the SV lens distribution over the range of refractive error where the deviation was less than 5%. Figures 5 and 6 show the mean number of lenses with 95% confidence intervals for the myopic and hyperopic tails of the ADD lens distribution. These are compared with the GHS over the same range of refractive error. The GHS was found to be statistically indistinguishable from the 1,000 generated distributions as it mostly sat within the 95% confidence intervals.

**Figure 3:**
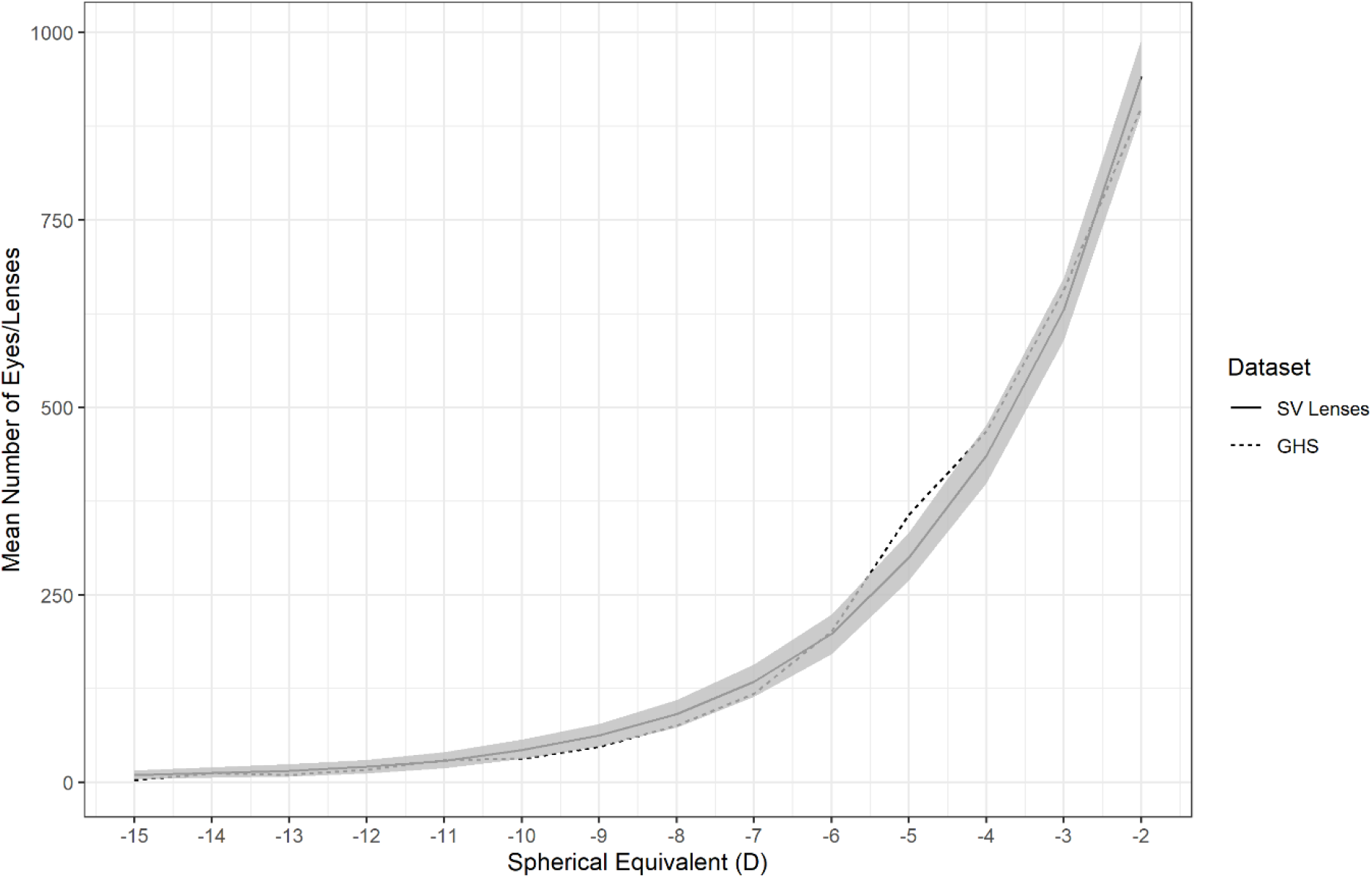
Bootstrapped myopic mean distribution with 95% confidence intervals for Single Vision lenses (solid line and shaded area) compared to the Gutenberg Health Survey (dotted line).

**Figure 4:**
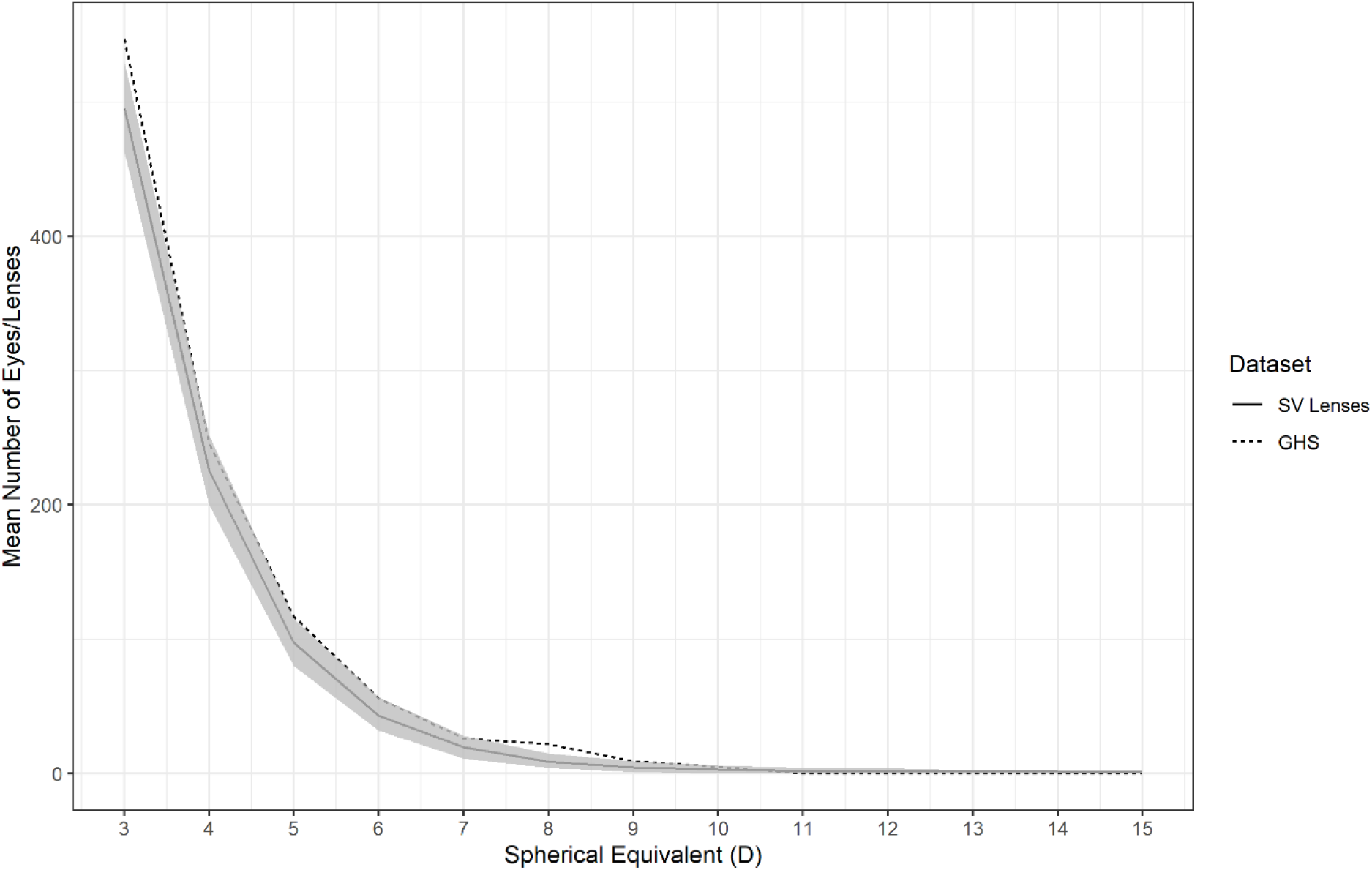
Bootstrapped hyperopic mean distribution with 95% confidence intervals for Single Vision lenses (solid line and shaded area) compared to the Gutenberg Health Survey (dotted line).

**Figure 5:**
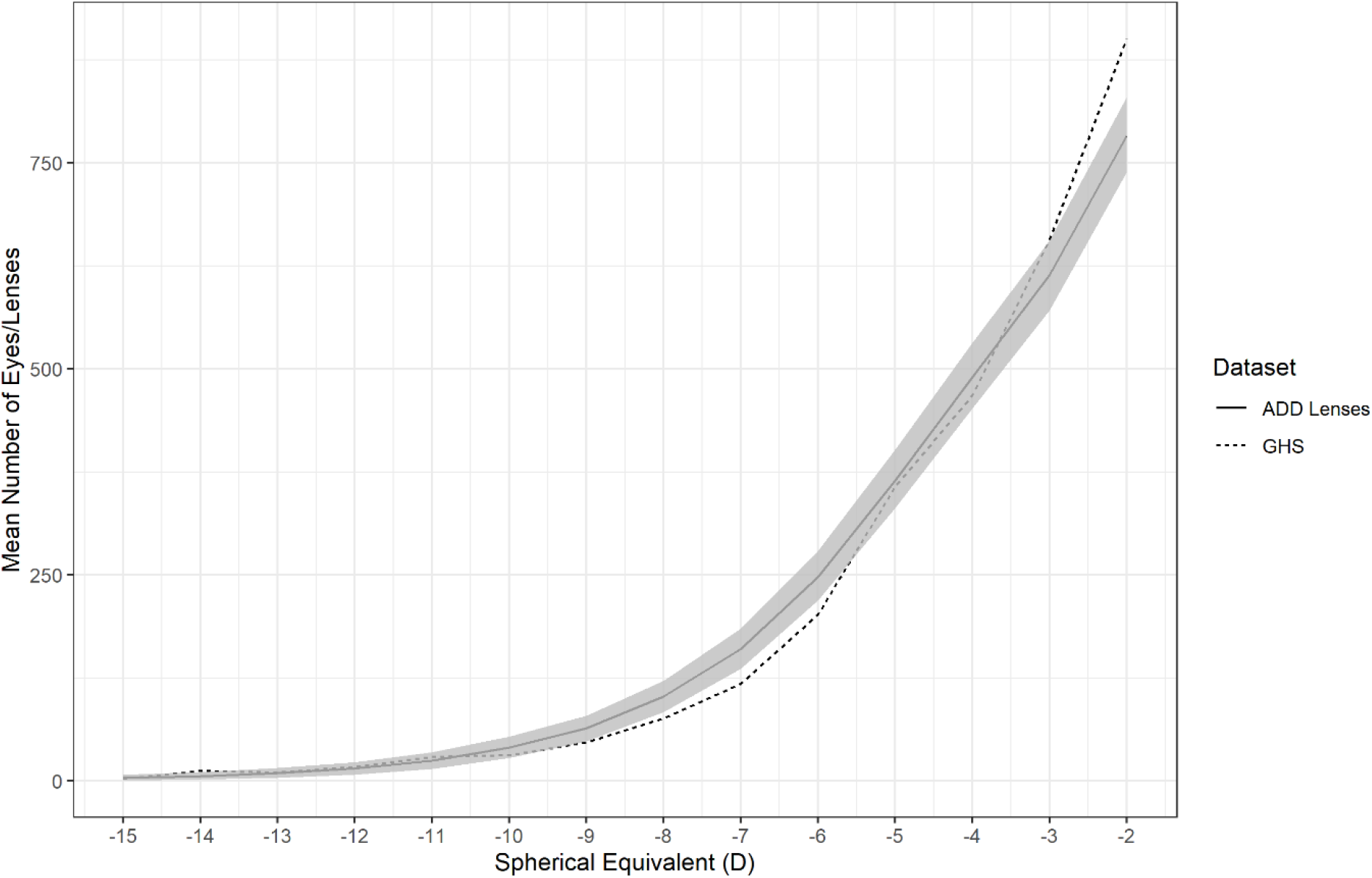
Bootstrapped myopic mean distribution with 95% confidence intervals for Addition lenses (solid line and shaded area) compared to the Gutenberg Health Survey (dotted line).

**Figure 6:**
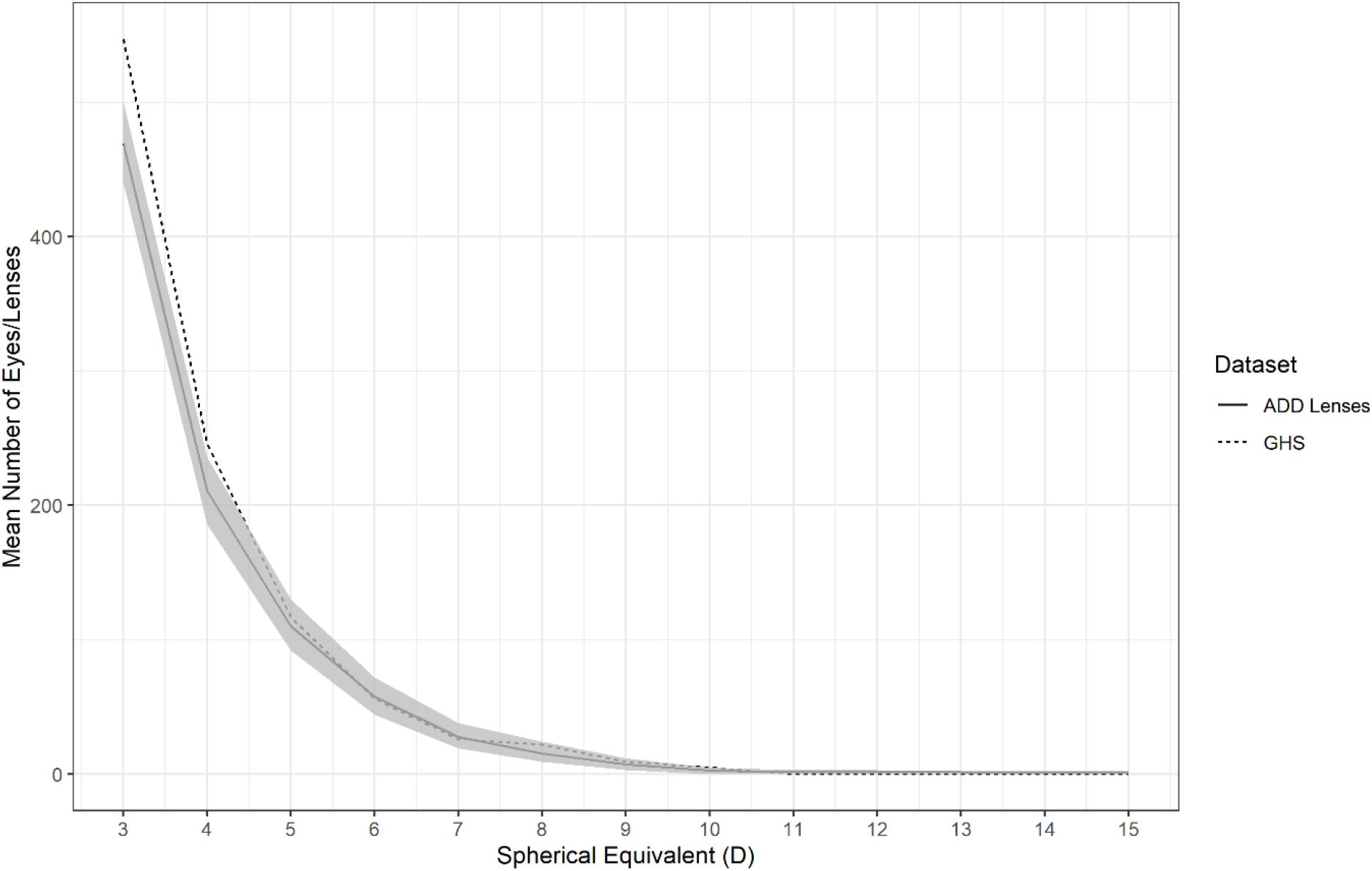
Bootstrapped hyperopic mean distribution with 95% confidence intervals for Addition lenses (solid line and shaded area) compared to the Gutenberg Health Survey (dotted line).

As the tails of the spectacle lens distributions were found to match the GHS between -2 D to -15D and +3 D to +15D, it was possible to determine the estimated risk of vision impairment at age 75 among myopic SV spectacle lens wearers (Table 2). Using the spectacle lens data, it was estimated that 9.58% of myopic spectacle lens wearers (n = 9,575 cases per 100,000 population) will be vision impaired by age 75. Over the same range of myopia in the GHS, 9.01% of myopic individuals (n = 9,012 cases per 100,000 population) were estimated to be vision impaired by age 75. The estimated rates of vision impairment were not statistically significantly different (χ^2^ = 182, DoF = 169, p = 0.234).

**Table 2:** Refractive distribution within the myopic tail of the SV spectacle lens data and the GHS data, estimated risk of vision impairment at age 75 from equation 1 and 2 and estimated number of individuals with vision impairment at age 75 per 100,000 people with myopia of -2 D or worse for both the SV lens group and GHS.

| Spherical Equivalent (D) | Proportion of myopia in SV lenses | Proportion of myopia in GHS | Risk of Vision Impairment at Age 75 | Estimated number of cases of vision impairment due to myopia per 100,000 population at age 75 (spectacle lens data) | Estimated number of cases of vision impairment due to myopia per 100,000 population at age 75 (GHS data) |
| --- | --- | --- | --- | --- | --- |
| -2 | 0.322 | 0.307 | 4% | 1,288 | 1,228 |
| -3 | 0.215 | 0.224 | 5% | 1,075 | 1,120 |
| -4 | 0.149 | 0.160 | 7% | 1,043 | 1,210 |
| -5 | 0.102 | 0.122 | 9% | 918 | 1,098 |
| -6 | 0.068 | 0.069 | 12% | 816 | 828 |
| -7 | 0.046 | 0.040 | 16% | 736 | 640 |
| -8 | 0.031 | 0.026 | 22% | 682 | 572 |
| -9 | 0.022 | 0.016 | 28% | 616 | 448 |
| -10 | 0.015 | 0.011 | 37% | 555 | 407 |
| -11 | 0.010 | 0.010 | 47% | 470 | 470 |
| -12 | 0.007 | 0.006 | 59% | 413 | 354 |
| -13 | 0.005 | 0.003 | 71% | 355 | 213 |
| -14 | 0.004 | 0.004 | 83% | 332 | 332 |
| -15 | 0.003 | 0.001 | 92% | 276 | 92 |
| **Total estimated vision impairment cases per 100,000 population at age 75 due to myopia ≤ -2.00D** |  |  |  | **9,575** | **9,012** |

## Discussion

This study describes a new method to estimate refractive error distribution. For spherical equivalent refractive errors exceeding +3 D for hyperopia and -2 D for myopia, spectacle lens sales data can provide equivalent estimates of the distribution of refractive error to those determined by conventional population surveys of refractive error. Furthermore, by accurately estimating the shape of the hyperopic and myopic tails of the distribution outside these threshold levels, this approach can provide useful estimates of future population risks of vision impairment.

There are some limitations with the use of spectacle lens sales data. Ametropes may not be corrected with spectacle lenses for numerous reasons, including, for example, lack of access to correction or the use of alternative forms of correction such as contact lenses. The majority of contact lens wearers also make use of spectacles, however, with only approximately 15% of contact lens wearers reporting they did not own any spectacles and over 75% reporting their spectacle prescription provided clear vision indicating it was up to date.^32^ It is also not possible to account for individuals who may have had refractive or cataract surgery in the current dataset. The literature indicates, however, that although the rates of surgery have increased they still represent less than 1% of all individuals.^33,34^ Given the very large sample size exploited herein, it is unlikely that these factors would have a significant impact on the results. This is supported by the similar levels of vision impairment predicted using both the spectacle lens data and GHS data. By utilising multiple datasets, it may be possible to better account for individuals not captured within spectacle lens data. In Europe, statistics are published on the number of surgical procedures performed,^35^ with similar data available for most countries,^36^ which may account for those undergoing cataract and refractive surgery. Applying the same methodology to contact lens sales data can account for patients that only use contact lenses for refractive error correction.

Other limitations also apply to the use of industrial type datasets. Drawing conclusions on subpopulations, for example, can be more difficult as spectacle lens manufacturers and other industry suppliers do not typically record data on their customers gender, ethnicity or age. It has, however, previously been demonstrated that lenses with a reading addition can be used to estimate a customer’s age.^25^ As the relationship between increasing age and increasing refractive error is the primary driver for vision impairment due to refractive error,^2,13,15^ accurate forecasting for the population risk of vision impairment using this methodology should be possible.

Emmetropes are also not well represented within this data. This is not surprising as it is unlikely that individuals with minimal or no refractive error purchase spectacle lenses in any significant quantities. This can be observed by the atypical distribution of refractive error for the SV lenses in Figure 1. It was expected that the ADD lens data would provide a closer match to the GHS in the emmetropic range due to the similar age profile to the GHS and the near universality of presbyopia over the age of 50.^25^ A likely explanation for the deviation of the ADD lens data at emmetropia in Figure 2 is the wide availability of over the counter reading glasses that can be used by emmetropes and low hyperopes and the ability of low myopes to read comfortably when no correction is in place, meaning those in this range of SE are less likely to purchase progressive addition spectacle lenses. The lack of representation of those with approximately emmetropic refractive errors in our data is a significant limitation, but epidemiological studies are best placed to establish baseline vision impairment risks for emmetropes/near emmetropes. In the GHS, the percentage of individuals estimated to be vision impaired by age 75 increases by 1.2% to 10.21% if those with myopia > -2.00 D SE are included in the calculations, which translates to approximately 10.81% if extrapolated to the SV spectacle lens wearers. Further modelling of the spectacle lens data may allow for more accurate estimates of the proportion of individuals in this low myopia group which in turn could allow a full population estimate of vision impairment due to myopia to be calculated. From a public health perspective, obtaining the current population burden of those with higher absolute refractive errors, especially myopia, is of particular importance as we are entering an era where myopia can reasonably be considered as a modifiable risk factor for vision impairment. These represent the individuals most at risk of vision impairment due to refractive error and estimating the number of people affected by higher refractive error can allow better public health planning.^37^

Due to the nature of the data, it is impossible to state how the refraction for each individual was carried out. Ideally all refractions would be carried out under cycloplegia in order to avoid the effects of accommodation, particularly for myopic refractions.^38^ It has been shown that the assessment of refractive error in adults is not significantly affected by the use of cycloplegia,^39^ particularly in older adults,^40^ those most at risk of vision impairment. The data used in this study likely represents predominantly adult populations, particularly the ADD lens data from which approximate ages can be calculated,^25^ so the probable lack of cycloplegia should have minimal effect on the refractive error and vision impairment estimations. It should also be noted that many well regarded population surveys of refractive error do not make use of cycloplegia^41,42^ including the main comparison study used herein.^27^ Additionally, the probable lack of cycloplegia in this study is unlikely to be significant as the higher myopic threshold should reduce the risk of a misclassification error and is the approach suggested by the International Myopia Institute when this risk may apply.^30^

The comparability of the results obtained from spectacle lens data and a conventional epidemiological study demonstrates the utility of industrial datasets as a public health tool in refractive error and vision impairment. The use of industrial data can potentially address the paucity of epidemiological data available for both refractive error^24^ and vision impairment.^43^ Manufacturers with large market share for spectacle lens sales may have refractive error data which can accurately determine the number of ametropes in a population and hence the risk of vision impairment due to refractive error, myopia in particular.

How this methodology could be best exploited to produce ongoing estimates of the population burden of refractive error and consequential vision impairment needs to be determined. The most significant challenge is gaining access to commercial data for public health purposes. One possible solution would involve the creation of an international consortium of industry, academic, professional, intergovernmental and non-governmental organisations and other key stakeholder bodies. This could provide a forum for international collaboration in the form of a Big Data coalition and lead to a Global Myopia Observatory of data analytic and data visualisation resources which could be used for public health planning, research, commercial and other uses. In providing the platform to gather and merge disparate sources of industry data, this consortium could provide a readily accessible, current and globally representative body of resources to monitor the changing epidemiology of refractive error and associated eye disease and the impact of new treatments and public health interventions essentially in real time. Furthermore, these resources would inform health planning decisions, drive clinical practice reform, stimulate industrial innovation and ultimately lead to better population health.

## Conclusion

The distribution of refractive error within a population over a large range of refractive error can be determined using spectacle lens sales data. This provides a good alternative when population level data on refractive error is either absent or outdated. This is a particularly useful methodology to determine the population burden of higher absolute levels of refractive error which represents the population cohort most at risk of vision impairment due to refractive error. An estimation of the future risk of vision impairment due to myopia can also be calculated from such data.

## Data Availability

The data contains potentially identifying or sensitive patient information. Access requests can be made to the TU Dublin Research Ethics and Integrity Committee.

